# Who is pregnant? defining real-world data-based pregnancy episodes in the National COVID Cohort Collaborative (N3C)

**DOI:** 10.1101/2022.08.04.22278439

**Authors:** Sara Jones, Katie R. Bradwell, Lauren E. Chan, Courtney Olson-Chen, Jessica Tarleton, Kenneth J. Wilkins, Qiuyuan Qin, Emily Groene Faherty, Yan Kwan Lau, Catherine Xie, Yu-Han Kao, Michael N. Liebman, Federico Mariona, Anup Challa, Li Li, Sarah J. Ratcliffe, Julie A. McMurry, Melissa A. Haendel, Rena C. Patel, Elaine L. Hill, the N3C Consortium

## Abstract

**Objective:** To define pregnancy episodes and estimate gestational aging within electronic health record (EHR) data from the National COVID Cohort Collaborative (N3C).

**Materials and Methods:** We developed a comprehensive approach, named <u>H</u>ierarchy and rule-based pregnancy episode <u>I</u>nference integrated with <u>P</u>regnancy <u>P</u>rogression <u>S</u>ignatures (HIPPS) and applied it to EHR data in the N3C from 1 January 2018 to 7 April 2022. HIPPS combines: 1) an extension of a previously published pregnancy episode algorithm, 2) a novel algorithm to detect gestational aging-specific signatures of a progressing pregnancy for further episode support, and 3) pregnancy start date inference. Clinicians performed validation of HIPPS on a subset of episodes. We then generated three types of pregnancy cohorts based on the level of precision for gestational aging and pregnancy outcomes for comparison of COVID-19 and other characteristics.

**Results:** We identified 628,165 pregnant persons with 816,471 pregnancy episodes, of which 52.3% were live births, 24.4% were other outcomes (stillbirth, ectopic pregnancy, spontaneous abortions), and 23.3% had unknown outcomes. We were able to estimate start dates within one week of precision for 431,173 (52.8%) episodes. 66,019 (8.1%) episodes had incident COVID-19 during pregnancy. Across varying COVID-19 cohorts, patient characteristics were generally similar though pregnancy outcomes differed.

**Discussion:** HIPPS provides support for pregnancy-related variables based on EHR data for researchers to define pregnancy cohorts. Our approach performed well based on clinician validation.

**Conclusion:** We have developed a novel and robust approach for inferring pregnancy episodes and gestational aging that addresses data inconsistency and missingness in EHR data.

## OBJECTIVE

Pregnancy episodes of care in EHR data are often inconsistently coded across patients. Our objective was to leverage pregnancy concepts recorded in the nationally-pooled EHR data of the National COVID Cohort Collaborative (N3C) to comprehensively infer pregnancy start and end dates using novel algorithms and to show the utility of these pregnancy episodes of care as a resource for COVID-19 research.

## BACKGROUND AND SIGNIFICANCE

The COVID-19 pandemic has substantially impacted daily life, and is especially concerning for vulnerable populations. Pregnant persons appear to be at higher risk for incident and severe COVID-19 infections than non-pregnant persons [1–4]. Increased rates of cesarean section deliveries, lower gestational age at delivery, and preterm birth have been observed among pregnant persons with COVID-19 [5]. Pregnant persons with COVID-19 may also present more frequently with acute respiratory distress syndrome and hemolysis, elevated liver enzymes, and low platelets syndrome [6, 7]. Moreover, access to and utilization of healthcare services markedly changed during the pandemic [1,2,8,9], which may have impacted antenatal care. Numerous knowledge gaps related to pregnancy and COVID-19 persist, including the impact of vaccination and the mechanisms underlying elevated risk for various poor outcomes.

EHR data can inform these knowledge gaps. The N3C [10] offers COVID-19 researchers access to harmonized EHR data from more than 12M individuals from 72 sites (7 April 2022). This is the first and largest publicly available EHR repository with national sampling in the U.S. However, data fields for start and end of pregnancies and gestational age at birth do not currently exist in a consistent form within EHRs, making it challenging to ascertain pregnancy episodes and gestational aging. Further, sites differ widely on how accurately and consistently this information is collected in their respective EHRs (Figure 1); moreover, the “gold-standard” of birth certificate-based data are usually unavailable in EHRs.

**Figure 1.**
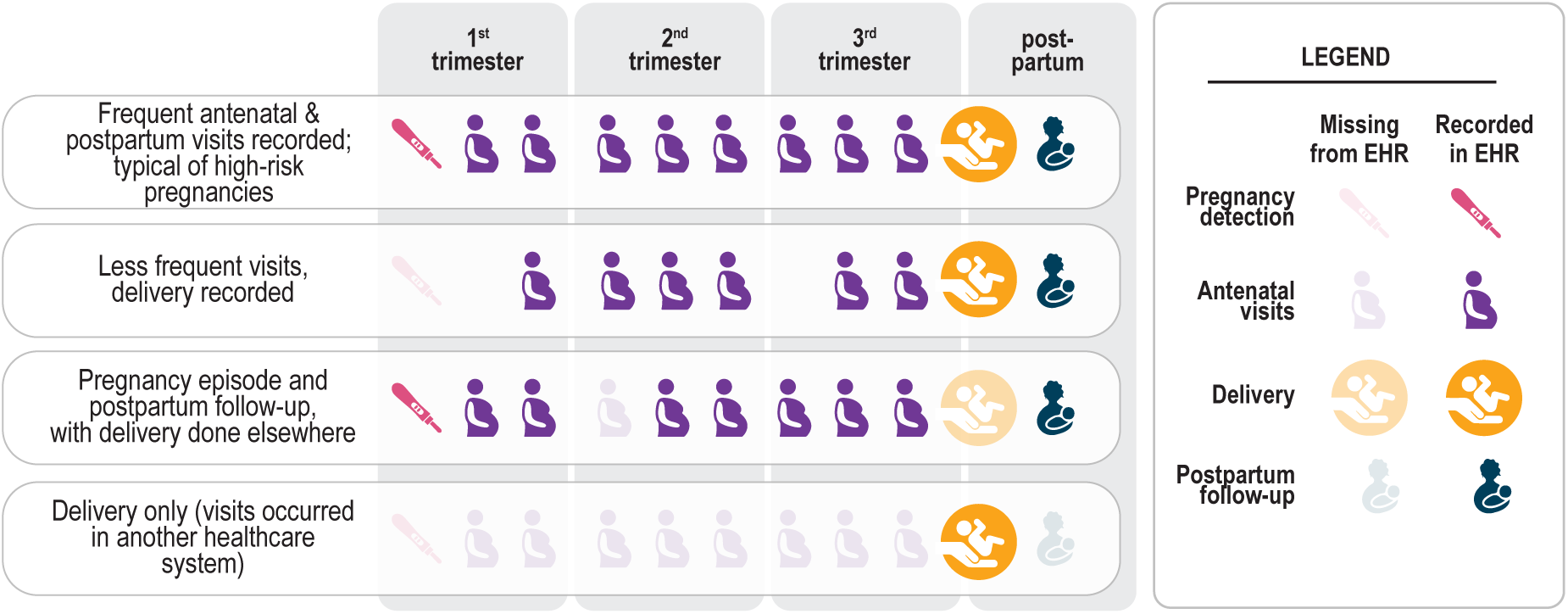
Common scenarios of pregnancy episodes in N3C illustrate how the EHR provides an incomplete picture of care. Some visits were not recorded (occurred in another healthcare system), and yet others were likely recorded inconsistently or inaccurately. Some routine visits may have occurred in another healthcare system or may not have occurred at all, potentially due to healthcare disruption caused by the pandemic.

To overcome these limitations, EHR-based investigations commonly utilize algorithms for determining gestational age and pregnancy (or delivery) episodes using diagnostic codes and delivery dates [11–15]. In N3C, we started with an existing algorithm using Observational Medical Outcomes Partnership (OMOP) concepts defined in Matcho et. al. [12]. However, this and similar algorithms rely heavily on anchoring the pregnancy with a specifically recorded pregnancy outcome (e.g. live birth), and do not leverage available pregnancy data for which an outcome is missing or may not have yet occurred. They also rely on approximations based on outcome date [16, 17] or hierarchies of a limited number of pregnancy markers [12] to infer pregnancy start, which may not always be recorded accurately in the data or may be absent from the EHR. Therefore we adapted Matcho et. al [12] and layered on own data-driven algorithms to robustly and precisely infer pregnancy start, pregnancy end, and landmark time frames throughout a pregnancy’s progression to provide temporal context.

## METHODS

### N3C cohort preparation

Detailed information on overall N3C organization can be found in previously published reports [10]. Briefly, each N3C data partner site provides demographic, visit, vital status, medication, laboratory, and diagnoses data; data is harmonized to the OMOP data model version 5.3.1 [18]. The N3C cohort is comprised of COVID-19-positive patients as well as COVID-19-negative controls matched on as many of the four sociodemographic variables (i.e., age, sex, race, and ethnicity) as possible. COVID-19-positive patients have been defined previously [19]. For patients included in N3C, encounters in the same data partner site on or after 1 January 2018 are also included (i.e., “lookback data”).

### Overview of our HIPPS approach to inferring pregnancy episodes, gestational aging, and pregnancy start dates

Our composite algorithm, named <u>H</u>ierarchy and rule-based pregnancy episode Inference integrated with <u>P</u>regnancy <u>P</u>rogression <u>S</u>ignatures (HIPPS), uses existing and data-driven analytic methods to identify pregnancies (Figure 2); HIPPS classifies each as a “pregnancy episode” recording all pregnancies per person (i.e., allowing an individual to be pregnant more than once during our observation period).

**Figure 2.**
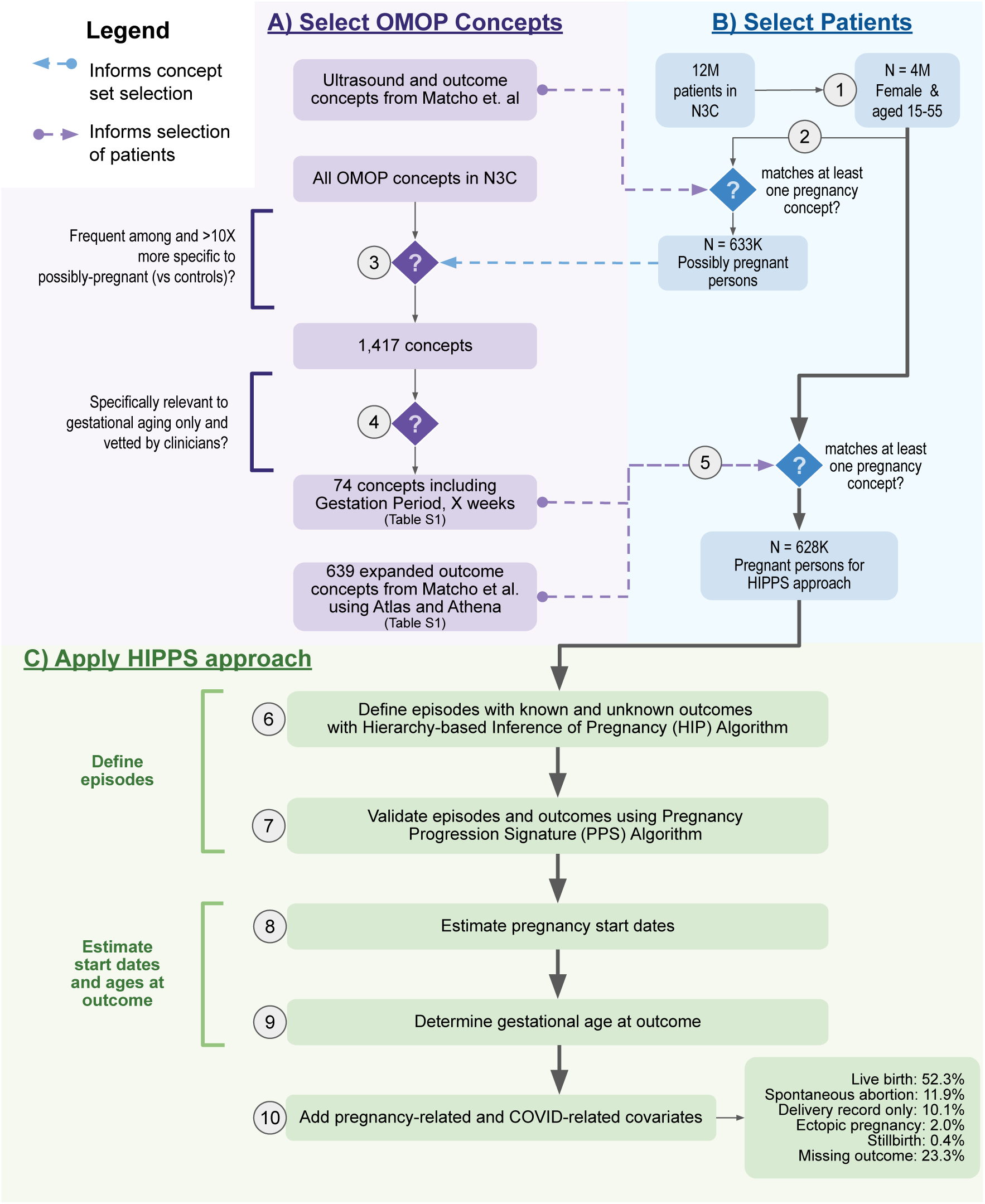
Overview of HIPPS. 1) From the 12M patients in N3C, we identified 4M that were both female and of reproductive age (15-55 years). 2) Of these, we identified 633K possibly pregnant persons who matched at least one concept in an initial set of ultrasound and pregnancy outcome concepts from Matcho et al. [12] 3) To develop an enriched set of concepts specific for pregnancy, we then assessed concept frequency among the initial cohort of possibly pregnant persons as compared with controls (all patients in N3C) and chose 1,417 concepts that were present in at least 1,000 individuals and were 10x (determined empirically via distribution analysis) more frequent among possibly pregnant persons. 4) Of 1,417 concepts, 74 concepts with fixed gestational time ranges <=3 months during pregnancy were vetted by clinicians and selected. 5) Using the 74 concepts and the 639 concepts expanded from Matcho et al. related to pregnancy outcomes, 628K pregnant persons were selected for the HIPPS approach [12]. 6) For each pregnant person, pregnancy episodes were defined with the Hierarchy-based Inference of Pregnancy (HIP) Algorithm. 7) Episodes and outcomes identified were validated by the Progressing Pregnancy Signature (PPS) Algorithm. 8) Pregnancy start dates were calculated using the Estimated Start Date (ESD) Algorithm. 9) Using the pregnancy start dates, gestational age in weeks was determined at the date of the pregnancy outcome. 10) Lastly, we added other pregnancy-related and COVID-related covariates to each episode.

Briefly, we first identified all females of reproductive age (15-55 years, inclusive) within N3C and applied pregnancy-specific concepts to identify potential pregnancies (Figure 2.1-5). Next, we applied two distinct algorithms to infer and validate unique pregnancy episodes (Figure 2.6-7). Then, we used gestational timing concepts to rigorously determine pregnancy episode start dates and their precision level (Figure 2.8-9). Finally, we merged the pregnancy episodes data with other pregnancy-related information as well as metrics important to COVID-19 research (Figure 2.10).

### Selecting relevant concepts for computable pregnancy phenotype and gestational aging

To define a computable phenotype for pregnancy in N3C, we started from a previously published concept set [12] and enriched it with additional concepts related to outcomes and gestational aging. This resulted in an expanded list of OMOP concepts that were relevant to either known pregnancy outcomes (639 concepts) or gestational aging (74 concepts) (Table S1).

### Hierarchy-based Inference of Pregnancy (HIP) Algorithm: Rule-based algorithm

To capture pregnancies without an outcome recorded in N3C, we developed the HIP Algorithm. Firstly, we used the “Outcome assessment and classification” step [12] to define episodes with the following outcomes: live birth, stillbirth, ectopic pregnancy, abortion (both spontanous and induced), and delivery record only. We then developed a novel approach to capture gestation-based episodes without outcomes, using gestational aging concepts that were prevalent in N3C data and had week-level resolution for estimating gestational age such as “Gestation Period, X weeks”. We calculated the start date of a gestation-based episode by tracking backwards the maximum gestational age in weeks from the end date. Lastly, gestation-based episodes were combined with outcome-based episodes if they overlapped.

### Pregnancy Progression Signature (PPS) Algorithm: Novel temporal sequence analysis for detecting pregnancy episodes

We developed the PPS algorithm to validate plausible episodes. PPS searches for signatures of progressing pregnancy concepts across each patient record using an adaptation of longest increasing consecutive subsequence analysis [20]. Briefly, time intervals across these concepts in the person’s data were compared to their expected intervals based on gestational ranges that clinicians assigned to the concepts (Figure 3A, Figure S1). Outcomes were added to each episode, and their concordance to HIP was assessed.

**Figure 3.**
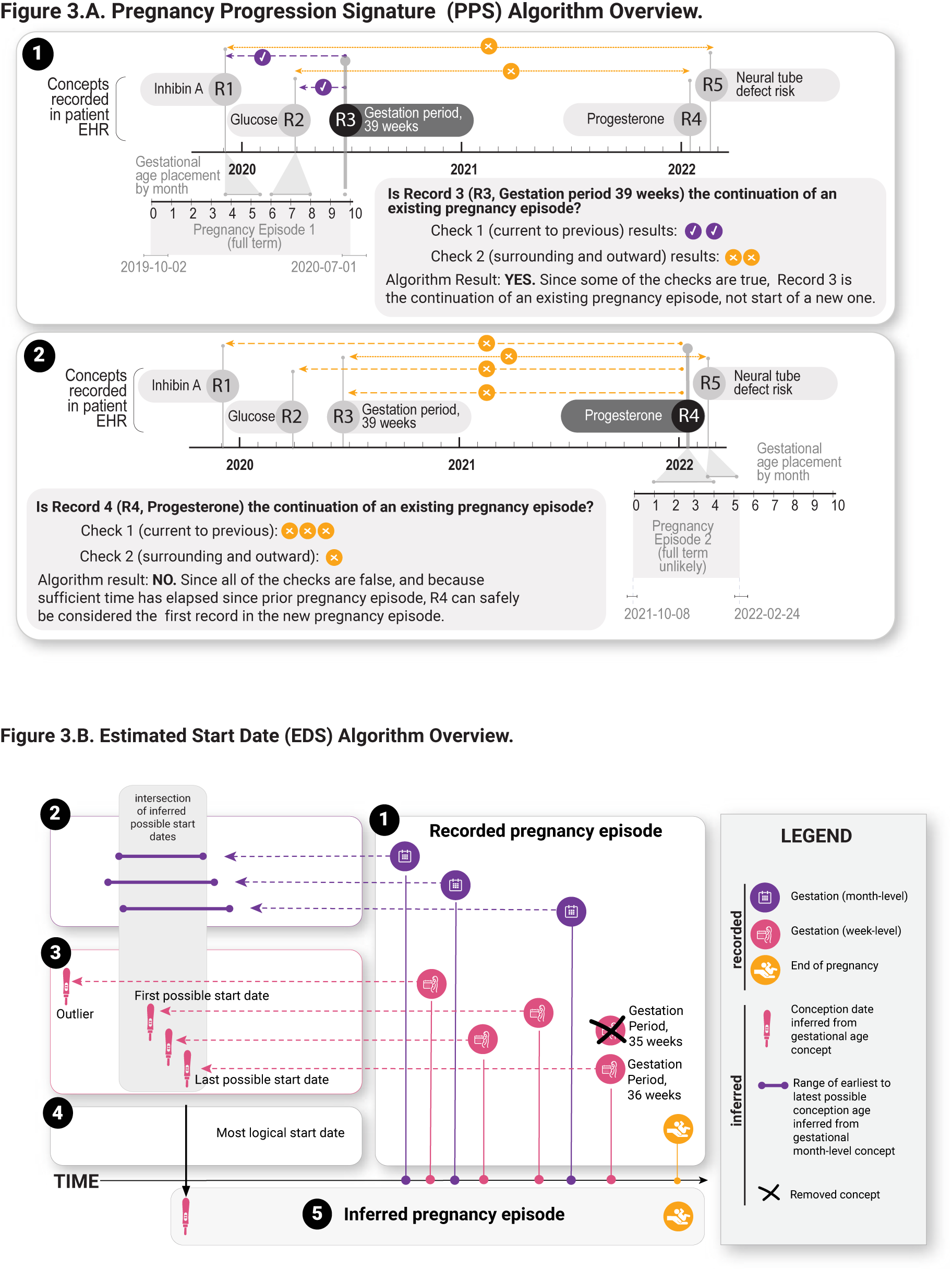
Overview of Pregnancy Progression Signature (PPS) Algorithm and Estimated Start Date (EDS) Algorithm. A) PPS Algorithm Overview. Notional examples of a continuing (1) and new (2) pregnancy episode are shown. Iteration and checks proceed for all records of a patient. In (1), PPS is iterating through the third record (R3) of the patient’s data. At R3, comparisons across concepts and record dates are performed via two main checks (Check 1, concept comparisons from current record to each previous record: dashed line, Check 2, concept comparisons surrounding and progressively outward from current record: dotted lines). Since at least one comparison evaluates to true (agreement between expected concept gestational timing and actual date differences between records), the episode is continued. In (2), the fourth patient record (R4) is reached in the same patient. As there are no checks that evaluate to true, a new episode is begun. B) Estimated Start Date Algorithm (EDSA) Overview. 1) Get all one-week to 3 month resolution (GR3m) and gestation week-level resolution (GW) concepts and their record dates during the recorded pregnancy episode. If there is more than one GW concept on the same record date, keep the max GW concept. 2) Get the min and max start ranges for each GR3m concept and find the intersection of these ranges. 3) Get the start dates of each GW concept. Any start date that falls outside of the intersection is considered an outlier. 4) Calculate the precision in days using the first and last start date in the intersection and set the last start date as the start of the inferred pregnancy episode (5).

### Estimated Start Date (ESD) Algorithm - Estimating precise pregnancy start dates

To estimate the start date for identified pregnancies (Figure 3B) we leveraged the set of 74 clinician-validated concepts, which included concepts with gestation week-level resolution (i.e., “GW” concepts like “Gestation Period, 12 weeks”) and resolution between one-week and 3 months (i.e., “GR3m” concepts like “Trisomy 18 risk [Likelihood] in Fetus,” which is generally assessed between 12 and 24 weeks). We filtered out outlier concepts, taking into account the timing of when each concept was recorded, and estimated the pregnancy start dates and corresponding levels of precision. GW concepts that occur latest in pregnancy were used to extrapolate the pregnancy start date based on empirical analysis (see Results). For comparison, we used a baseline method that obtains the start dates using only the GW concepts within an episode without any removal of outliers and assigned precision based on the maximum start date difference between GW concepts.

### Addition of pregnancy-related information to each pregnancy episode

For each pregnant person and pregnancy episode, we ascertained available sociodemographic information (e.g., age, race/ethnicity) and other pregnancy-related information e.g., type of delivery (vaginal vs. cesarean), number of fetuses (singleton vs. multiple), and preterm status. Although parity and gravidity are not directly ascertainable in N3C (due to the limited observation period relative to each pregnant person’s lifespan), HIPPS itself is well suited to address these questions in other datasets. We created OMOP concept sets (Table S1 and Table S2) for pregnancy-related variables from all possible concepts available in various OMOP concept databases (e.g., Athena [21] and Atlas [22], Figure 2.4); all concept sets were vetted by clinicians on our team.

### Validation by clinicians and independent pregnancy-specific concepts

Manual chart review is not possible in N3C due to regulations that minimize reidentification risk; we therefore provided two obstetrician-gynecologists with an N3C dataset containing all OMOP concepts related to measurements, observations, conditions, drugs, and procedures for each of 60 randomly-selected pregnancy episodes from 46 pregnant persons, of whom 13 had more than one recorded pregnancy (Figure S2): (1) 50 pregnancy episodes consisting of four pregnancy outcomes to yield 10 episodes per outcome/group and 20 for live birth, (2) 10 pregnancy episodes without outcomes, or (3) no pregnancy episodes identified. Within the group with no pregnancy episodes identified, we randomly selected 10 females of reproductive age with a pregnancy-related concept yet no evidence of pregnancy progression. Each clinician then assessed the presence, number, dates, and outcomes of each pregnancy episode as well as the gestational age in weeks at outcome and estimated last menstrual period (LMP). Additionally, the clinicians rated their level of confidence in annotating the above information with categories of not confident (-1), neutral (0), or confident (1). We computed the percent agreement for pregnancy outcome category, gestational age in weeks, date-related metrics, and the total number of episodes between each of the 60 instances assessed by both HIPPS and the clinicians’ validations, to quantify concordance of HIPPS with clinicians’ validations. Additionally, to provide further validation for pregnancy episode completeness and gestational timing, we specifically checked overlap and timing within pregnancy episodes of an independent set of 25 concepts (Table S3). These concepts were validated by clinicians to occur exclusively during pregnancy over a range wider than was suitable for episode definition by our algorithms (four to 10 months).

### COVID-19 analysis

We used descriptive statistics to compare demographic characteristics across three types of pregnancy cohorts based on pregnancy episodes: (1) all episodes regardless of outcome category (i.e., a least conservative definition of a pregnancy cohort) (**cohort 1: all episodes**), (2) episodes with outcomes (but excluding delivery record only) and pregnancy start dates with up to one month resolution for estimated start date of pregnancy (includes week level poor support; i.e., a moderately conservative definition of a pregnancy cohort) (**cohort 2: month-level)**, and (3) pregnancy episodes with outcomes (but excluding delivery record only) with high concordance (score of 2) between HIP and PPS and down to week-level resolution for estimated start date of pregnancy (i.e., a most conservative definition of a pregnancy cohort) (**cohort 3: week-level)**. For each cohort, we further stratified pregnancy episodes into: (1) all episodes regardless of date or COVID-19 diagnosis status, (2) episodes ending before 1 March 2020, (3) episodes with indication of COVID-19-positivity after 1 March 2020, and (4) episodes with no indication of COVID-19-positivity status after 1 March 2020. We took the first occurrence of either a positive COVID-19 PCR or antigen (Ag) lab result or COVID-19 diagnostic code (U07.1) to assess COVID-19 positivity during a person’s pregnancy. Among pregnant persons who were COVID-19-positive during their pregnancy, we examined N3C COVID-19 severity measures [23].

## RESULTS

To identify pregnancy episodes we used 713 OMOP concepts, including 74 concepts for inferring pregnancy start dates (Figure 2A). As of data extraction date (7 April 2022), 72 unique data partner sites contributed data for >12M total patients in N3C. Among these patients, HIPPS identified 4M females of reproductive age (15-55), of which a further subset of 628K had at least one pregnancy episode recorded in N3C (Figure 2B). We identified 816,471 episodes of which 426,852 (52.3%) had live births, 3,545 (0.4%) stillbirths, 97,506 (11.9%) spontaneous abortions, 16,014 (2.0%) ectopic pregnancies, 82,201 (10.1%) with delivery record only, and 190,353 (23.3%) missing outcome recorded (Figure 2C). Of those with pregnancy episodes, 51.4% are White non-Hispanic, 17.7% are Hispanic or Latino any race, 19.1% Black/African American non-Hispanic, 4.3% Asian American non-Hispanic, 0.2% Native Hawaiian or Pacific Islander non-Hispanic, 6.2% unknown, and 1.1% other non-Hispanic.

### Gestational timing concept curation and exploration

Details of the data availability analysis and other validation analyses can be found in the Supplement (Figure S3-5). For each of the 1,417 pregnancy-specific concepts we determined the mean month and standard deviation of when the concept occurs relative to the pregnancy start of HIP-inferred episodes with outcomes (Figure S6). We retained a total of 145 concepts after filtering out concepts with a standard deviation >1.5 months (Table S3). Clinicians assigned expected gestational time ranges to each of the 145 concepts (Table S3); we kept the 74 concepts with time ranges ≤ 3 months. The vast majority of records of the 43 clinician-curated GR3m gestational timing concepts occurred during the expected time ranges using HIP’s episodes (Figure 4). Similarly, the records of the GW concepts aligned with their corresponding clinician gestational time range.

**Figure 4.**
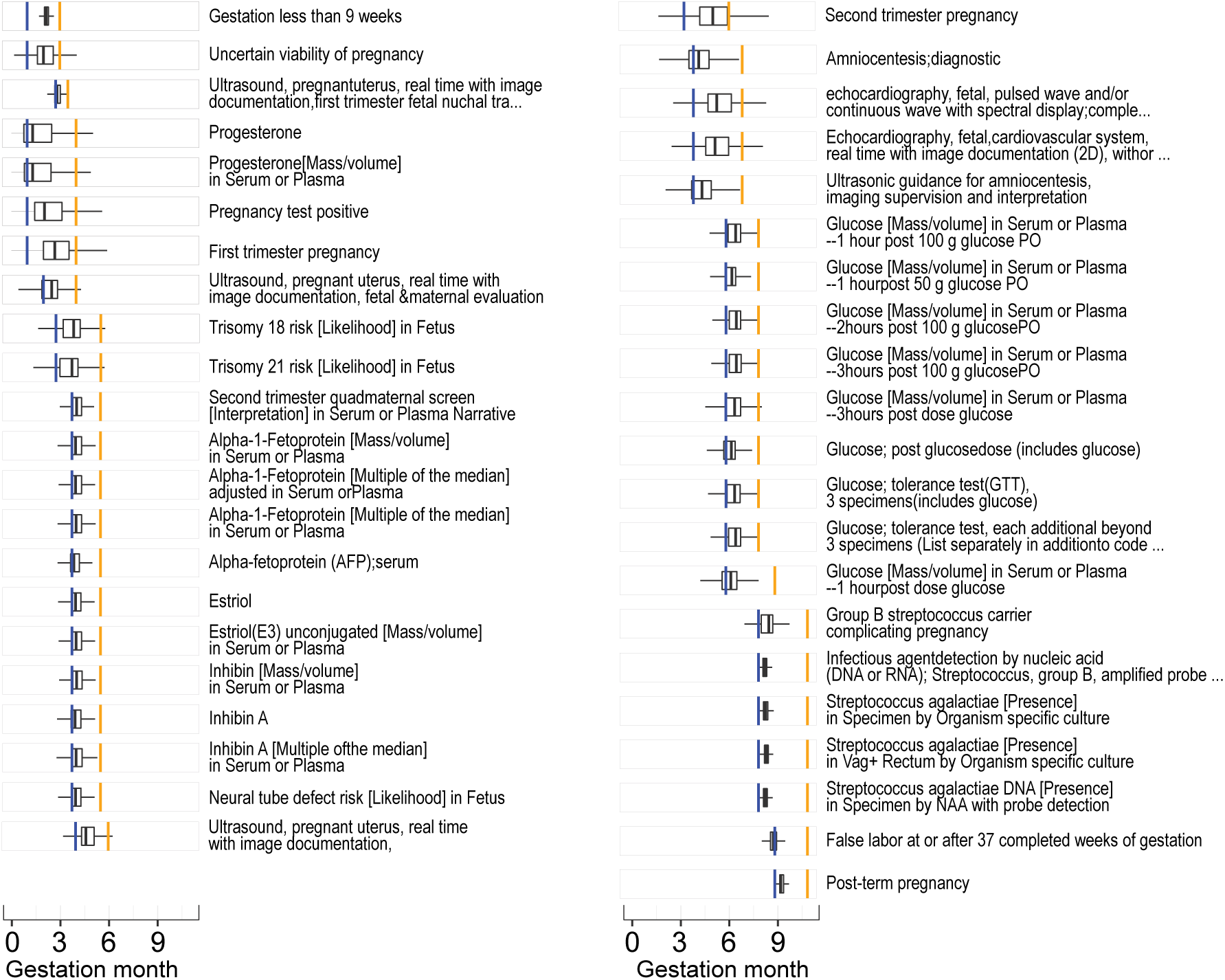
Gestational timing precision for clinician-curated concepts of > 1 week and ≤ 3 months (GR3m). Agreement between clinician-assigned gestation ranges (blue bars = min, orange bar = max) and IQR of concepts in the data (boxplots) for the concepts selected as input for algorithm 2 with clinician gestational time ranges > 1 week and ≤ 3 months. For simplicity, we have not shown the Gestation period, X weeks (GW) concepts as these are all narrow by definition.

### Pregnancy start date estimation and evaluation of precision

For those episodes with GW concepts, we selected start dates with GW concepts with higher gestational timing reliability (Figure S6). Using the GW and GR3m concepts for ESD, our algorithm yielded 431,173 (52.8%) episodes with week-level (>1 concept) resolution. Compared with a baseline method, our algorithm achieved a 41.0% increase in episodes with week-level precision and reduced the number of episodes for non-specific precision by 49.1% (Table S4). Across all episodes with pregnancy outcomes, the proportion of episodes with week-level resolution increased (Figure 5A, Table S5).

**Figure 5.**
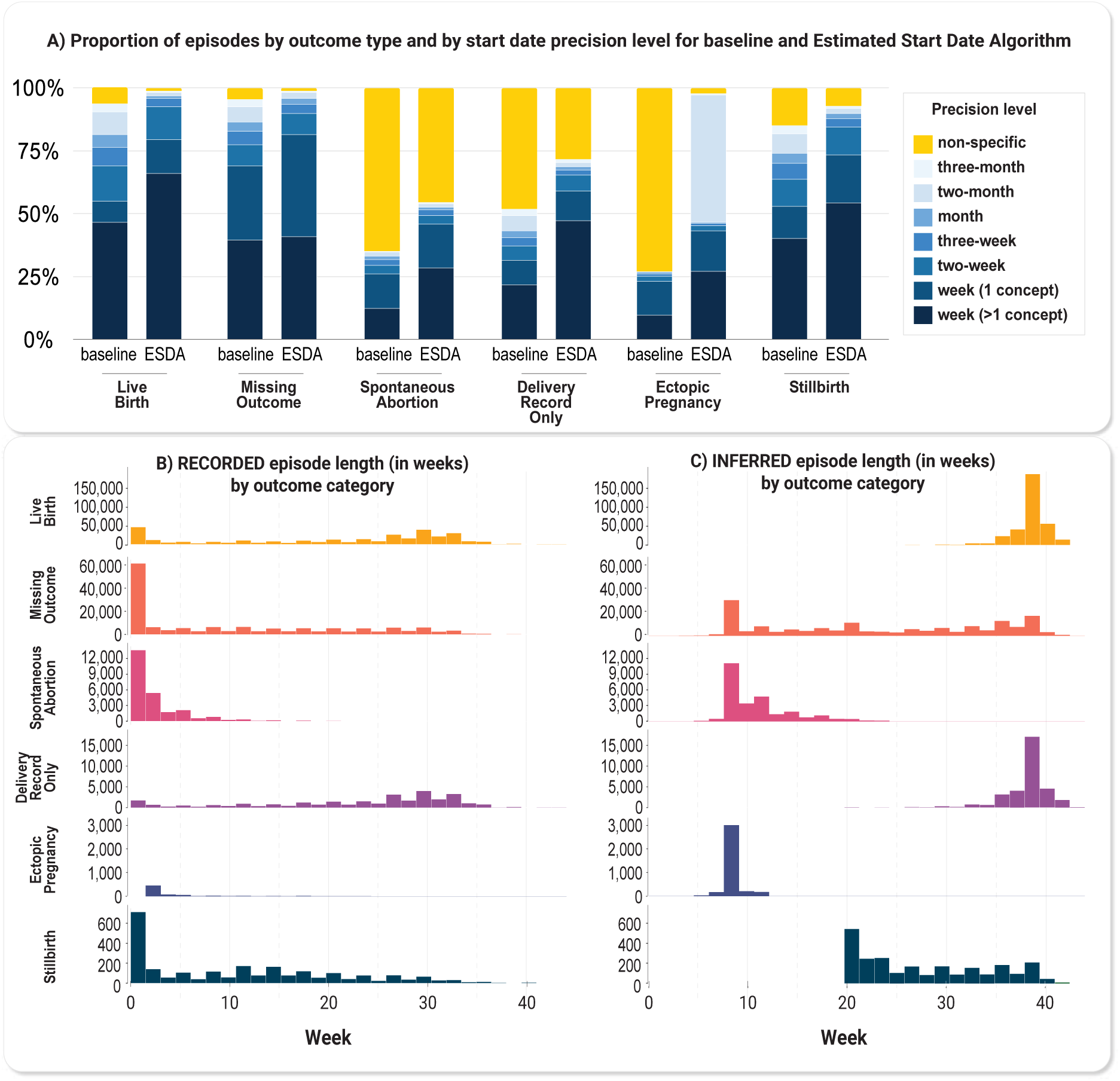
HIPPS results. A) Proportion of episodes by outcome category and by start date precision level for baseline and Estimated Start Date Algorithm. The baseline method obtained the start dates using only the GW concepts within an episode without any removal of outliers and assigned precision based on the maximum start date difference between GW concepts. B) Histogram of episodes with week-level resolution only (N=564,762) by outcome category of recorded pregnancy lengths (start and end dates of records for pregnancy that occur within the EHR data) in weeks and C) inferred pregnancy lengths (actual pregnancy start and end estimated using HIPPS) in weeks.

### Characterization of full set of episodes from HIPPS approach

Pregnancy episodes identified by our HIPPS approach found a significant proportion of likely ongoing pregnancies, had appropriate pregnancy lengths for respective outcome categories, where the vast majority of episodes identified by HIP algorithm were also supported by PPS, and common outcomes had a high concordance score. Of the 190,353 (23.3%) of the episodes missing an outcome, 41,629 (21.9%) of these likely represent an ongoing pregnancy as these episodes had not reached the time range of plausible delivery outcomes for, for example, live birth or stillbirth (Figure S7A). Over 40.0% of episodes across all outcomes that occur after 20 weeks of gestation have only one outcome concept within 28 days after pregnancy outcome date, suggesting these may be delivery-only type records (Figure S7B). Our algorithmically-inferred pregnancy length corresponded well with the expected time intervals by outcome category (Figure 5C). Of the episodes first inferred by HIP, 724,251 (88.7%) episodes were supported by PPS. Of those not supported by PPS (92,220 episodes, 11.3%), 74,493 (80.8%) HIP episodes did not have any gestational information and 81,151 (88.0%) episodes only had a recorded length of one day. When looking at the outcome concordance score, a metric of support for the identified outcomes, both live birth and stillbirth tended to have a higher percentage of episodes with the highest score of 2 (Figure S8). Spontaneous abortions and ectopic pregnancies on the other hand tended to have a higher percentage of episodes with the lowest score of 0.

### Clinician validation

Clinician validation agreed 100.0% with HIPPS-identified episodes (Cohen’s kappa coefficient 0.8591 and asymptotic 95% CI, 0.7628 to 0.9553; exact p-value of test for chance agreement: 4.87×10^-33^) (Table 1). The clinicians identified three extra episodes among the 13 pregnant persons with multiple episodes (one with no outcome and two with spontaneous abortions), thus agreement was assessed among a total of 63 instances: 63 episodes considered by clinicians (as ‘gold standard’) without regard for randomly-sampled 10 instances assessed by algorithm as being without episodes (Table 1).

**Table 1.** Independent internal validation of episodes and outcome categories by clinicians.

|  | Number of episodes identified by algorithm | Number of episodes identified by clinician | Percent completeness compared to clinician review | Percent agreement between algorithm and clinician | Clinicians' average confidence score | Standard deviation of clinicians' confidence scores |
| --- | --- | --- | --- | --- | --- | --- |
| **Persons without episodes** | 10 | 10 | 100.0 | 100.0 | N/A | N/A |
| **Number of episodes (among 46 females)** | 60 | 63 | 100.0 | 100.0 | N/A | N/A |
| **Outcome** |  |  |  |  |  |  |
| Live birth | 20 | 21 | 95.2 | 100.0** | 0.9 | 0.31 |
| Stillbirth | 10 | 9 | 100.0* | 90.0 | 0.89 | 0.33 |
| Spontaneous Abortion | 10 | 14 | 71.4 | 100.0** | 1 | 0 |
| Ectopic Pregnancy | 10 | 9 | 100.0* | 90.0 | 0.89 | 0.33 |
| **Gestational age at outcome ($\pm 1$ week)** |  |  |  |  |  |  |
| Live birth | N/A | N/A |  | 100.0 | 0.6 | 0.50 |
| Stillbirth | N/A | N/A |  | 87.5 | 0.5 | 0.53 |
| Spontaneous Abortion | N/A | N/A |  | 77.8 | -0.44 | 0.52 |
| Ectopic Pregnancy | N/A | N/A |  | 77.8 | -0.44 | 0.52 |

\* All of the clinical curation-defined episodes for these categories were verified by algorithmic assessment.
\*\* All of the algorithmically-defined episodes for these categories were verified by clinical assessment.
Note: As these cases were randomly selected for clinician validation they do not reveal information about the subpopulation and are not subject to the same policy regarding obfuscating small numbers of patients.

The percent agreement by outcome category, including no outcome, was over 80.0% and the average confidence score ranged from 0.89 to 1.00. Within one week, HIPPS had high agreement with clinicians’ validations: 78.0% to 100.0% for start dates, 89.0% to 100.0% for end dates, and 78.0% to 100.0% for gestational age. These average confidence scores tended to be higher for live birth vs. other outcomes (Table 2).

**Table 2.**
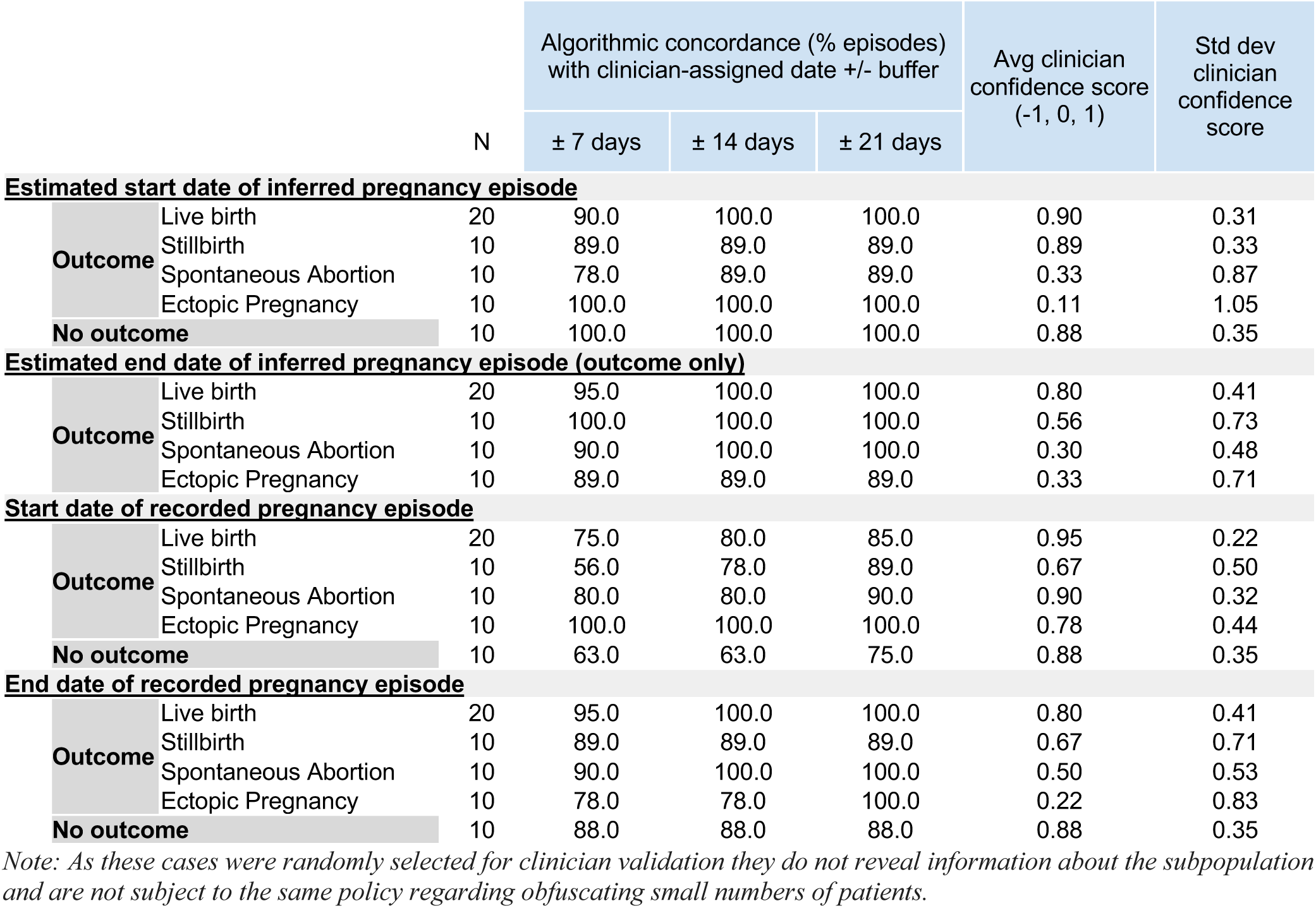
Independent internal validation of start and end dates of both inferred and recorded pregnancy episodes by clinicians.

Similarly, our additional check using the 25 clinician-validated concepts, that were independent to our algorithm definition and known to occur specifically *within* pregnancy, indicated a mean of 90.2 (SD = 5.4)% of the concepts overlapped with the expected gestational time ranges for episodes (Table S6).

### Characterization of the various pregnancy cohorts stratified by COVID-19 status

Among the approximate 816,471 pregnancy episodes (cohort 1 all episodes), 489,776 (60.0%) had month-level granularity for gestational start date (cohort 2: month-level) and 271,044 (33.2%) had week-level granularity for gestational start date (cohort 3: week-level) (Table S7). Comparing across cohorts, patient characteristics were fairly similar with a few exceptions: patients in cohort 3 (week-level) were less likely to be 15-17 years old, were more likely to have a defined outcome of live birth, and more likely to have COVID-19 screening during pregnancy (Figure 6). There were minimal differences in patient characteristics before versus after March 2020 (Figure 6).

**Figure 6.**
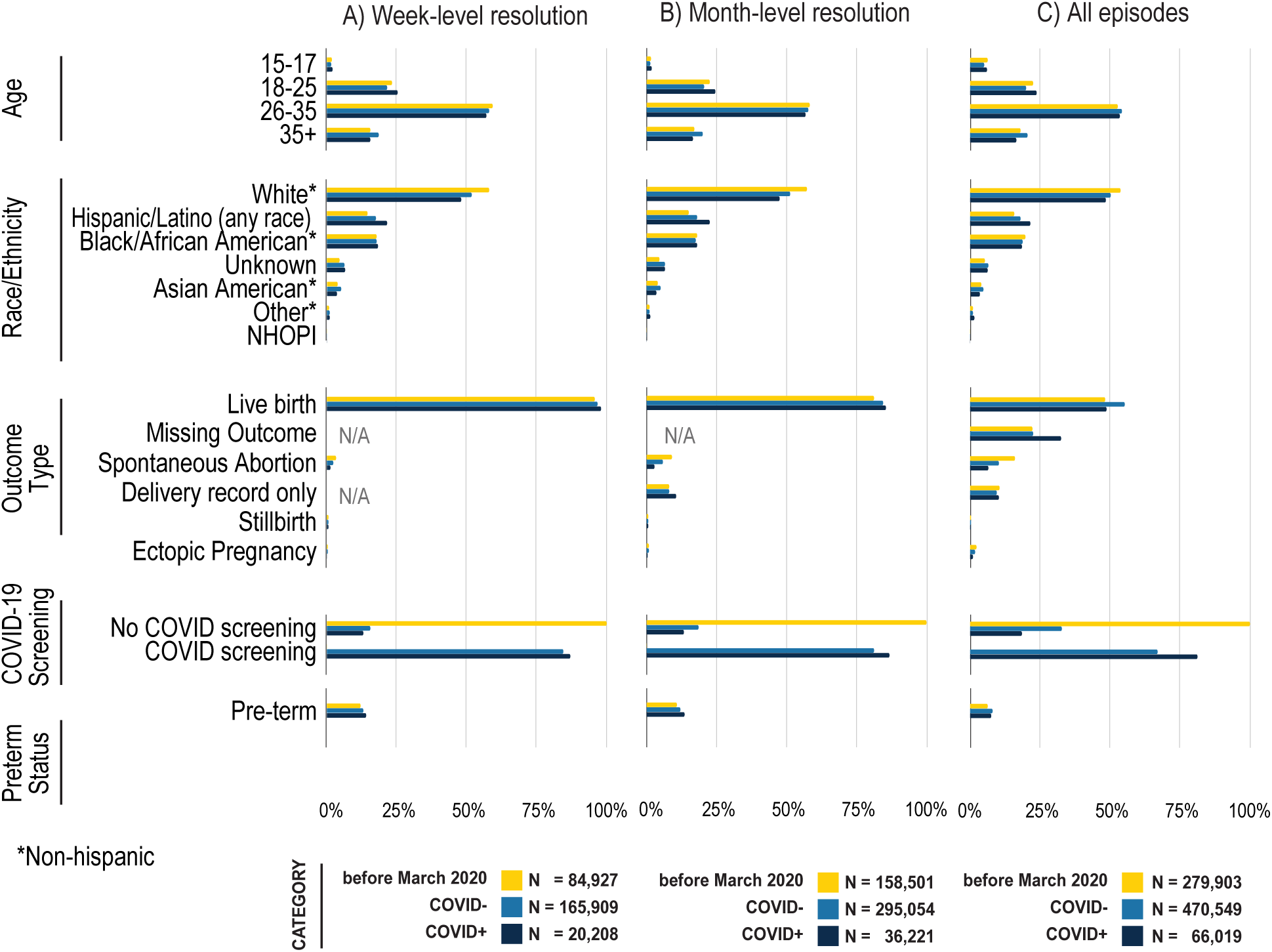
Demographics and outcomes of pregnant persons before and during COVID-19 pandemic, stratified by week-level resolution (Panel A), month-level resolution (Panel B), and all patients (Panel C). See Table S7 for source data. Note that COVID negative (COVID-) includes pregnant persons without any results in their records.

For episodes post-March 2020, we further stratified by COVID-19 positivity during pregnancy; COVID- 19-positive pregnant persons were more likely to be Hispanic (21.5% vs. 18.2%), more likely to be 18-25 years old and less likely to be 35+ years old. Those with a positive test or diagnosis were more likely to be screened for COVID-19 than those without (81.3% versus 67.1% Table S7). These trends were similar for cohorts 2 and 3, however, missing data was less frequent among these cohorts compared to cohort 1, possibly indicating a more complete EHR record.

For all episodes with a positive COVID-19 test during pregnancy, we explored demographic characteristics by COVID-19 severity (Table 3).

**Table 3.** Demographics and outcomes of pregnant persons by COVID-19 severity and reinfection status.

|  | Mild No ED* or Hosp around COVID index |  | Mild ED* around COVID index |  | Moderate Hosp around COVID index |  | Severe ECMO IMV* in Hosp around COVID index |  | Mortality after COVID index |  | Reinfection |  |
| --- | --- | --- | --- | --- | --- | --- | --- | --- | --- | --- | --- | --- |
|  | N | % | N | % | N | % | N | % | N | % | N | % |
| **Age** |  |  |  |  |  |  |  |  |  |  |  |  |
| 15-17 | 3,018 | 6.4 | 258 | 5.3 | 666 | 5.1 | < 20 | < 12.3 | < 20 | < 31.7 | < 20 | < 2.0 |
| 18-25 | 10,257 | 21.9 | 1,508 | 31.2 | 3,660 | 28.1 | 40 | 24.5 | < 20 | < 31.7 | 258 | 25.2 |
| 26-35 | 25,555 | 54.5 | 2,383 | 49.4 | 6,758 | 51.8 | 84 | 51.5 | 35 | 55.6 | 597 | 58.3 |
| 35+ | 8,083 | 17.2 | 679 | 14.1 | 1,953 | 15.0 | 35 | 21.5 | < 20 | < 31.7 | 162 | 15.8 |
| **Race/Ethnicity** |  |  |  |  |  |  |  |  |  |  |  |  |
| White Non-Hispanic | 24,684 | 52.6 | 1,873 | 38.8 | 4,808 | 36.9 | 77 | 47.2 | 25 | 39.7 | 529 | 51.7 |
| Hispanic or Latino Any Race | 9,425 | 20.1 | 1,053 | 21.8 | 3,509 | 26.9 | 33 | 20.2 | < 20 | < 5.4 | 188 | 18.4 |
| Black/African American Non-Hispanic | 7,391 | 15.8 | 1,460 | 30.2 | 3,171 | 24.3 | 27 | 16.6 | 21 | 33.3 | 233 | 22.8 |
| Unknown | 3,019 | 6.4 | 257 | 5.3 | 903 | 6.9 | < 20 | < 12.3 | < 20 | < 5.4 | 38 | 3.7 |
| Asian American Non-Hispanic | 1,587 | 3.4 | 119 | 2.5 | 427 | 3.3 | < 20 | < 12.3 | < 20 | < 5.4 | 27 | 2.6 |
| Other Non-Hispanic | 710 | 1.5 | 53 | 1.1 | 199 | 1.5 | < 20 | < 12.3 | < 20 | < 5.4 | < 20 | < 1.9 |
| NHOPI* Non-Hispanic | 97 | 0.2 | < 20 | < 0.4 | 20 | 0.2 | < 20 | < 12.3 | < 20 | < 5.4 | < 20 | < 1.9 |
| **Preterm Status** |  |  |  |  |  |  |  |  |  |  |  |  |
| Yes | 2,974 | 6.3 | 226 | 4.7 | 1,769 | 13.6 | 78 | 47.9 | < 20 | < 19.0 | 41 | 4 |
| **Total\*\*** | 46,913 | 71.1 | 4,828 | 7.3 | 13,037 | 19.7 | 163 | 0.2 | 63 | 0.1 | 1,024 | 1.6 |

Highlighted in yellow are values that were slightly adjusted to obfuscate any back calculations of patient numbers under 20.
\*NHOPI = Native Hawaiian or Other Pacific Islander, ED = Emergency Department, ECMO = extracorporeal membrane oxygenation, IMV = invasive mechanical ventilation
\*\*Percentage is out of 66,019 episodes with COVID-19 positivity.

Among 66,019 (8.1%) episodes with COVID-19 positivity, 71.1% of the infections were mild, 7.3% involved an emergency department visit, 19.7% resulted in a hospitalization, less than 1.0% required extracorporeal membrane oxygenation (ECMO) or resulted in mortality, and 1.6% were a reinfection. Black/African American non-Hispanic and Hispanic and older pregnant persons experienced greater severity compared to their counterparts. Preterm birth was markedly higher amongst those with severe vs. moderate COVID-19 (47.9% vs. 13.6%).

## DISCUSSION

We have not only created the largest cohort to date of COVID-19 positive pregnant persons from the U.S. but we have also developed a comprehensive, novel resource for others to precisely infer pregnancy episodes in any EHR data. In this geographically and demographically diverse and historically unprecedented U.S. dataset, we have identified 816,471 pregnancy episodes, with the majority of episodes having week-level resolution for pregnancy start. Our work provides a strong foundation for follow-up work for researchers and programs looking to address major knowledge gaps regarding pregnancy and COVID-19, and beyond.

Our composite algorithm, HIPPS, combines two independent algorithms, each developed to address the disjointed or noncontinuous EHR records of a pregnant person: one algorithm, HIP, is an extension of a previously-developed rule-based algorithm [12] and the other is PPS, an entirely novel algorithm based on the longest increasing consecutive subsequence analysis. We used PPS to validate episodes from HIP. HIPPS offers unique elements not currently covered by existing, published algorithms. First, since the best prior algorithm only considers pregnancies with an outcome (e.g. live birth), pregnancies without such records are omitted by definition [12]. As pregnancies without outcome may offer important data, particularly related to COVID-19 research, both our HIP and PPS algorithms were designed to capture pregnancies with and without outcomes, the former by employing “Gestation Period, X weeks” concepts as attempted by Naleway et al. [15]. Second, PPS can check if the gestational timing data is progressing, even without outcome anchoring. Third, our HIPPS approach is consistent with how clinicians would annotate EHR information as demonstrated by our clinician validation. Fourth, high concordance between HIP and PPS provides high confidence in our pregnancy episode and outcome inference. Fifth, combining empirical analysis and clinician validation to derive gestational timing concepts for PPS and ESD allows episode inference to be optimized to the EHR dataset available. This is an advantage over using an inflexible set of concepts of previous algorithms [12, 15] for inferring pregnancy episode timing that may not be generalizable to other EHR datasets.

Our final novel sub-algorithm applied to the episodes, ESD, rigorously estimates pregnancy start date; this is ever more important in ascertaining risks related to exposures in pregnancy and outcomes, including for COVID-19. By including a validation metric, precision in days, for ESD, we provide a high level of confidence for the estimated start date of a pregnancy, with over 52.8% and 67.3% episodes with week- and month-level resolution, respectively. Such precision in gestational aging allows researchers to determine associations with an unprecedented level of confidence.

Finally, as proof-of-concept of application of HIPPS to N3C data, we generated three types of cohorts of pregnant persons, ranging from least-conservatively defined (all episodes regardless of precision in gestational aging) to most-conservatively defined cohorts (week-level precision for gestational age and defined outcome), and compared sociodemographic, clinical, and COVID-19 variables across the cohorts. While we generally did not observe many significant differences in the proportion of pregnant persons, some interesting differences arose among the three types of pregnancy cohorts we presented. For example, those in the most conservatively defined cohort are less likely to be teenaged, more likely to have live birth outcomes, and more likely to have been screened for COVID-19 than those in our other two cohorts. That this cohort type is less likely to have spontaneous abortion or ectopic pregnancy is only natural given these outcomes have less gestational info. Thus, for example, questions anchoring on vaccination timing during pregnancy on risk of breakthrough or incident COVID-19 infection may require this most conservative cohort of pregnant persons, but then evaluating spontaneous abortions as the outcome would be a likely mismatch. On the other hand, certain research questions may not require exact gestational aging, as is common with claims data when researchers assign 40 weeks gestation to classify exposure where gestational info is missing, in which case selecting the least conservative cohort may be most appropriate. Our presentation of three possible cohorts, to elucidate the possible sizes, demographics, precision, and tradeoffs for using exact definition types to define pregnancy cohorts in N3C, should aid future researchers for various research efforts not only within N3C but also for other EHR-based studies.

While our work has several unique strengths, limitations also exist. First, we are unable to conduct source validation. Due to the nature of N3C making only de-identified data available that cannot be re-identified, we cannot conduct a “gold standard” validation of our findings against source data using birth certificates or medical charts; thus our work, in principle, cannot ascertain accuracy. Though we attempted a clinician validation approach that mimicked a medical chart review, it is possible that both the clinician annotators and HIPPS misidentified a pregnancy episode or misclassified a pregnancy outcome given the fundamental similar source of both. Regardless, previous literature supports our approach for defining pregnancy episodes even without source data validation [12]. Second, as with any EHR dataset, the likelihood of missingness and misclassification is high. It is possible that we may have missed potential pregnancy episodes or outcomes with our current concept sets, although we found inferred episode overlap of 90.2% on average between the independent 25 gestation age-specific concepts to those used for algorithm definition. This suggests that concept bias, either in our overall algorithms or in the small numbers included in our clinician validation, is unlikely. Also, because we did not filter out any episodes with a single occurrence of an outcome concept, as Matcho et al. had done [12], it is possible that our dataset overestimates the presence of some outcomes. Relatedly, misclassification may be high too, especially for uncommon pregnancy outcomes, and depending on the research question this could create sources of bias. Nonetheless, since we do provide the number of outcome concepts and confidence score for each pregnancy episode, users of the data and our approach can individualize criteria for excluding episodes depending on their research question. Third, while our approach has extremely high precision on inferences for common outcomes, such as live birth, we observed less precision in inferences related to less common outcomes (i.e. ectopic pregnancies), or underreported outcomes, (i.e. spontaneous abortions), which are common issues among all EHR-based phenotyping of such outcomes.

## CONCLUSION

Using HIPPS to identify pregnancy episodes and outcomes, we have not only developed the largest cohort to date of COVID-19 positive pregnant persons from the U.S. but enabled gestational aging with high precision-crucial to research attempting to ascertain associations between certain pregnancy-related exposures and outcomes, elements that are ever-more important in COVID-19-related research in pregnancy. Our novel approach to gestational aging has enduring implications for others to precisely infer pregnancy episodes in EHR data.

## ETHICS STATEMENT

Data partner sites transfer their N3C-eligible data to NCATS/NIH under a Johns Hopkins University Reliance Protocol (IRB00249128) or via individual site agreements with NCATS (see below). Managed under the NIH authority, the N3C Data Enclave can be accessed as previously described [10] and at ncats.nih.gov/n3c/resources, https://covid.cd2h.org/for-researchers.

| Site | IRB Name | Exempted vs approved | Protocol number |
| --- | --- | --- | --- |
| Medical University of South Carolina | Health Sciences South Carolina Institutional Review Board | exempt | Pro00111335 |
| National Institutes of Health | NIH Office of IRB Operations | exempt | N/A |
| University of Minnesota | University of Minnesota Institutional Review Board | approved | STUDY00012706 |
| University of Rochester | University of Rochester Research Subjects Review Board | exempt | STUDY00005366 |
| University of Washington | Human Subjects Division | approved | STUDY00013147 |

## CODE AVAILABILITY

All analyses were conducted in the N3C Enclave using R (v.3.5.1), Python (v.3.6.10), and PySpark (v.3.0.2). To support reproducibility, the code for implementing HIPPS, and full package list with versions is available to users with valid login credentials in the N3C enclave.

## DISCLAIMER

The content is solely the responsibility of the authors and does not necessarily represent the official views of the NIH.

Individual authors were supported by the following funding sources: NIMH R01131542 (RCP). The funding sources or study sponsors had no role in study design; in the collection, analysis, and interpretation of data; in the writing of the report; and in the decision to submit the paper for publication. SEJ was supported by an appointment to the National Institute of Allergy and Infectious Diseases (NIAID) Emerging Leaders in Data Science Research Participation Program. This program is administered by the Oak Ridge Institute for Science and Education through an interagency agreement between the U.S. Department of Energy (DOE) and NIAID. ORISE is managed by ORAU under DOE contract number DE-SC0014664. All opinions expressed in this paper are the authors’ and do not necessarily reflect the policies and views of NIAID, DOE, or ORAU/ORISE.

## Author Contributions

SEJ, KRB, RCP, and ELH designed the study.

SEJ and KRB prepared code utilized in obtaining the pregnancy episodes dataset and validation.

SEJ, KRB, and QQ analyzed the data.

KJW performed statistical analysis.

SEJ, KRB, JAM, EGF, LEC, and YKL prepared tables and figures.

CO-C, FM, and JT reviewed concepts.

CO-C and JT performed independent clinician validation of episodes.

SEJ, KRB, JAM, LEC, KJW, RCP, and ELH prepared the manuscript.

All authors reviewed the manuscript and provided feedback.

## Data Availability

N3C Data Enclave can be accessed at ncats.nih.gov/n3c/resources, https://covid.cd2h.org/for-researchers

https://covid.cd2h.org/for-researchers

## Acknowledgements

We would like to thank Caroline Signore for providing clinical expertise, Davera Gabriel for assisting in initial concept set creation, and the Logic Liaison Team for creating the logic and data for defining COVID positivity and severity. This work was supported by the National Institutes of Health, National Center for Advancing Translational Sciences Institute grant number U24TR002306.

The analyses described in this publication were conducted with data or tools accessed through the NCATS N3C Data Enclave covid.cd2h.org/enclave and supported by NCATS U24 TR002306. This research was possible because of the patients whose information is included within the data from participating organizations (covid.cd2h.org/dtas) and the organizations and scientists (covid.cd2h.org/duas) who have contributed to the on-going development of this community resource (cite this https://doi.org/10.1093/jamia/ocaa196).

## We gratefully acknowledge the following core contributors to N3C

Adam B. Wilcox, Adam M. Lee, Alexis Graves, Alfred (Jerrod) Anzalone, Amin Manna, Amit Saha, Amy Olex, Andrea Zhou, Andrew E. Williams, Andrew Southerland, Andrew T. Girvin, Anita Walden, Anjali A. Sharathkumar, Benjamin Amor, Benjamin Bates, Brian Hendricks, Brijesh Patel, Caleb Alexander, Carolyn Bramante, Cavin Ward-Caviness, Charisse Madlock-Brown, Christine Suver, Christopher Chute, Christopher Dillon, Chunlei Wu, Clare Schmitt, Cliff Takemoto, Dan Housman, Davera Gabriel, David A. Eichmann, Diego Mazzotti, Don Brown, Eilis Boudreau, Elizabeth Zampino, Emily Carlson Marti, Emily R. Pfaff, Evan French, Farrukh M Koraishy, Federico Mariona, Fred Prior, George Sokos, Greg Martin, Harold Lehmann, Heidi Spratt, Hemalkumar Mehta, Hongfang Liu, Hythem Sidky, J.W. Awori Hayanga, Jami Pincavitch, Jaylyn Clark, Jeremy Richard Harper, Jessica Islam, Jin Ge, Joel Gagnier, Joel H. Saltz, Johanna Loomba, John Buse, Jomol Mathew, Joni L. Rutter, Justin Starren, Karen Crowley, Katie Rebecca Bradwell, Kellie M. Walters, Ken Wilkins, Kenneth R. Gersing, Kenrick Dwain Cato, Kimberly Murray, Kristin Kostka, Lavance Northington, Lee Allan Pyles, Leonie Misquitta, Lesley Cottrell, Lili Portilla, Mariam Deacy, Mark M. Bissell, Marshall Clark, Mary Emmett, Mary Morrison Saltz, Matvey B. Palchuk, Meredith Adams, Meredith Temple-O’Connor, Michael G. Kurilla, Michele Morris, Nabeel Qureshi, Nasia Safdar, Nicole Garbarini, Noha Sharafeldin, Ofer Sadan, Patricia A. Francis, Penny Wung Burgoon, Peter Robinson, Philip R.O. Payne, Rafael Fuentes, Randeep Jawa, Rebecca Erwin-Cohen, Rena Patel, Richard A. Moffitt, Richard L. Zhu, Rishi Kamaleswaran, Robert Hurley, Robert T. Miller, Saiju Pyarajan, Sam G. Michael, Samuel Bozzette, Sandeep Mallipattu, Satyanarayana Vedula, Scott Chapman, Shawn T. O’Neil, Soko Setoguchi, Stephanie S. Hong, Steve Johnson, Tellen D. Bennett, Tiffany Callahan, Umit Topaloglu, Usman Sheikh, Valery Gordon, Vignesh Subbian, Warren A. Kibbe, Wenndy Hernandez, Will Beasley, Will Cooper, William Hillegass, Xiaohan Tanner Zhang. Details of contributions available at covid.cd2h.org/core-contributors

The following institutions whose data is released or pending:

Available: Advocate Health Care Network — UL1TR002389: The Institute for Translational Medicine (ITM) • Boston University Medical Campus — UL1TR001430: Boston University Clinical and Translational Science Institute • Brown University — U54GM115677: Advance Clinical Translational Research (Advance-CTR) • Carilion Clinic — UL1TR003015: iTHRIV Integrated Translational health Research Institute of Virginia • Charleston Area Medical Center — U54GM104942: West Virginia Clinical and Translational Science Institute (WVCTSI) • Children’s Hospital Colorado — UL1TR002535: Colorado Clinical and Translational Sciences Institute • Columbia University Irving Medical Center — UL1TR001873: Irving Institute for Clinical and Translational Research • Duke University — UL1TR002553: Duke Clinical and Translational Science Institute • George Washington Children’s Research Institute — UL1TR001876: Clinical and Translational Science Institute at Children’s National (CTSA-CN) • George Washington University — UL1TR001876: Clinical and Translational Science Institute at Children’s National (CTSA-CN) • Indiana University School of Medicine — UL1TR002529: Indiana Clinical and Translational Science Institute • Johns Hopkins University — UL1TR003098: Johns Hopkins Institute for Clinical and Translational Research • Loyola Medicine — Loyola University Medical Center • Loyola University Medical Center — UL1TR002389: The Institute for Translational Medicine (ITM) • Maine Medical Center — U54GM115516: Northern New England Clinical & Translational Research (NNE-CTR) Network • Massachusetts General Brigham — UL1TR002541: Harvard Catalyst • Mayo Clinic Rochester — UL1TR002377: Mayo Clinic Center for Clinical and Translational Science (CCaTS) • Medical University of South Carolina — UL1TR001450: South Carolina Clinical & Translational Research Institute (SCTR) • Montefiore Medical Center — UL1TR002556: Institute for Clinical and Translational Research at Einstein and Montefiore • Nemours — U54GM104941: Delaware CTR ACCEL Program • NorthShore University HealthSystem — UL1TR002389: The Institute for Translational Medicine (ITM) • Northwestern University at Chicago — UL1TR001422: Northwestern University Clinical and Translational Science Institute (NUCATS) • OCHIN — INV-018455: Bill and Melinda Gates Foundation grant to Sage Bionetworks • Oregon Health & Science University — UL1TR002369: Oregon Clinical and Translational Research Institute • Penn State Health Milton S. Hershey Medical Center — UL1TR002014: Penn State Clinical and Translational Science Institute • Rush University Medical Center — UL1TR002389: The Institute for Translational Medicine (ITM) • Rutgers, The State University of New Jersey — UL1TR003017: New Jersey Alliance for Clinical and Translational Science • Stony Brook University — U24TR002306 • The Ohio State University — UL1TR002733: Center for Clinical and Translational Science • The State University of New York at Buffalo — UL1TR001412: Clinical and Translational Science Institute • The University of Chicago — UL1TR002389: The Institute for Translational Medicine (ITM) • The University of Iowa — UL1TR002537: Institute for Clinical and Translational Science • The University of Miami Leonard M. Miller School of Medicine — UL1TR002736: University of Miami Clinical and Translational Science Institute • The University of Michigan at Ann Arbor — UL1TR002240: Michigan Institute for Clinical and Health Research • The University of Texas Health Science Center at Houston — UL1TR003167: Center for Clinical and Translational Sciences (CCTS) • The University of Texas Medical Branch at Galveston — UL1TR001439: The Institute for Translational Sciences • The University of Utah — UL1TR002538: Uhealth Center for Clinical and Translational Science • Tufts Medical Center — UL1TR002544: Tufts Clinical and Translational Science Institute • Tulane University — UL1TR003096: Center for Clinical and Translational Science • University Medical Center New Orleans — U54GM104940: Louisiana Clinical and Translational Science (LA CaTS) Center • University of Alabama at Birmingham — UL1TR003096: Center for Clinical and Translational Science • University of Arkansas for Medical Sciences — UL1TR003107: UAMS Translational Research Institute • University of Cincinnati — UL1TR001425: Center for Clinical and Translational Science and Training • University of Colorado Denver, Anschutz Medical Campus — UL1TR002535: Colorado Clinical and Translational Sciences Institute • University of Illinois at Chicago — UL1TR002003: UIC Center for Clinical and Translational Science • University of Kansas Medical Center — UL1TR002366: Frontiers: University of Kansas Clinical and Translational Science Institute • University of Kentucky — UL1TR001998: UK Center for Clinical and Translational Science • University of Massachusetts Medical School Worcester — UL1TR001453: The UMass Center for Clinical and Translational Science (UMCCTS) • University of Minnesota — UL1TR002494: Clinical and Translational Science Institute • University of Mississippi Medical Center — U54GM115428: Mississippi Center for Clinical and Translational Research (CCTR) • University of Nebraska Medical Center — U54GM115458: Great Plains IDeA-Clinical & Translational Research • University of North Carolina at Chapel Hill — UL1TR002489: North Carolina Translational and Clinical Science Institute • University of Oklahoma Health Sciences Center — U54GM104938: Oklahoma Clinical and Translational Science Institute (OCTSI) • University of Rochester — UL1TR002001: UR Clinical & Translational Science Institute • University of Southern California — UL1TR001855: The Southern California Clinical and Translational Science Institute (SC CTSI) • University of Vermont — U54GM115516: Northern New England Clinical & Translational Research (NNE-CTR) Network • University of Virginia — UL1TR003015: iTHRIV Integrated Translational health Research Institute of Virginia • University of Washington — UL1TR002319: Institute of Translational Health Sciences • University of Wisconsin-Madison — UL1TR002373: UW Institute for Clinical and Translational Research • Vanderbilt University Medical Center — UL1TR002243: Vanderbilt Institute for Clinical and Translational Research • Virginia Commonwealth University — UL1TR002649: C. Kenneth and Dianne Wright Center for Clinical and Translational Research • Wake Forest University Health Sciences — UL1TR001420: Wake Forest Clinical and Translational Science Institute • Washington University in St. Louis — UL1TR002345: Institute of Clinical and Translational Sciences • Weill Medical College of Cornell University — UL1TR002384: Weill Cornell Medicine Clinical and Translational Science Center • West Virginia University — U54GM104942: West Virginia Clinical and Translational Science Institute (WVCTSI)

Submitted: Icahn School of Medicine at Mount Sinai — UL1TR001433: ConduITS Institute for Translational Sciences • The University of Texas Health Science Center at Tyler — UL1TR003167: Center for Clinical and Translational Sciences (CCTS) • University of California, Davis — UL1TR001860: UCDavis Health Clinical and Translational Science Center • University of California, Irvine — UL1TR001414: The UC Irvine Institute for Clinical and Translational Science (ICTS) • University of California, Los Angeles — UL1TR001881: UCLA Clinical Translational Science Institute • University of California, San Diego — UL1TR001442: Altman Clinical and Translational Research Institute • University of California, San Francisco — UL1TR001872: UCSF Clinical and Translational Science Institute Pending: Arkansas Children’s Hospital — UL1TR003107: UAMS Translational Research Institute • Baylor College of Medicine — None (Voluntary) • Children’s Hospital of Philadelphia — UL1TR001878: Institute for Translational Medicine and Therapeutics • Cincinnati Children’s Hospital Medical Center — UL1TR001425: Center for Clinical and Translational Science and Training • Emory University — UL1TR002378: Georgia Clinical and Translational Science Alliance • HonorHealth — None (Voluntary) • Loyola University Chicago — UL1TR002389: The Institute for Translational Medicine (ITM) • Medical College of Wisconsin — UL1TR001436: Clinical and Translational Science Institute of Southeast Wisconsin • MedStar Health Research Institute — UL1TR001409: The Georgetown-Howard Universities Center for Clinical and Translational Science (GHUCCTS) • MetroHealth — None (Voluntary) • Montana State University — U54GM115371: American Indian/Alaska Native CTR • NYU Langone Medical Center — UL1TR001445: Langone Health’s Clinical and Translational Science Institute • Ochsner Medical Center — U54GM104940: Louisiana Clinical and Translational Science (LA CaTS) Center • Regenstrief Institute — UL1TR002529: Indiana Clinical and Translational Science Institute • Sanford Research — None (Voluntary) • Stanford University — UL1TR003142: Spectrum: The Stanford Center for Clinical and Translational Research and Education • The Rockefeller University — UL1TR001866: Center for Clinical and Translational Science • The Scripps Research Institute — UL1TR002550: Scripps Research Translational Institute • University of Florida — UL1TR001427: UF Clinical and Translational Science Institute • University of New Mexico Health Sciences Center — UL1TR001449: University of New Mexico Clinical and Translational Science Center • University of Texas Health Science Center at San Antonio — UL1TR002645: Institute for Integration of Medicine and Science • Yale New Haven Hospital — UL1TR001863: Yale Center for Clinical Investigation

