## Supplementary Figures for "Who is pregnant? defining real-world data-based pregnancy episodes in the National COVID Cohort Collaborative (N3C)"

1 **Get expected time ranges for each gestational-timing concept**

| Record | Concept date (A) | Concept | Min month (E) | Max month (E) |
|---|---|---|---|---|
| R1 | 2019-12-01 | Inhibin A [Multiple of the median] in Serum or Plasma | 3.75 | 5.5 |
| R2 | 2020-03-29 | Glucose; post glucose dose (includes glucose) | 6 | 8 |
| R3 | 2020-06-02 | Gestation period, 11 weeks | 2.75 | 2.75 |
| R4 | 2020-07-01 | Gestation period, 39 weeks | 9.75 | 9.75 |
| R5 | 2022-01-13 | Progesterone [Mass/volume] in Serum or Plasma | 1 | 4 |
| R6 | 2022-02-24 | Neural tube defect risk [Likelihood] in Fetus | 3.75 | 5.5 |

Discordant record (R3)

2 **Compare actual (*A*) vs. expected (*E*) time differences across records**

Check 1: Compare the current record (*i*) to all its previous (*i - x*) records

$$\left(E_i^{min} - E_{i-x}^{max}\right) - 2 \le \left(A_i - A_{i-x}\right)/30 \le \left(E_i^{max} - E_{i-x}^{min}\right) + 2$$

| *i* | *x* | *i - x* | Evaluated Equation | Result |
|---|---|---|---|---|
| R2 | 1 | R1 | -1.50 ≤ 3.97 ≤ 6.25 | True |
| R3 | 1 | R2 | -7.25 ≤ 2.17 ≤ -1.25 | False |
| R3 | 2 | R1 | -4.75 ≤ 6.13 ≤ 1.00 | False |
| R4 | 1 | R3 | 5.00 ≤ 0.97 ≤ 9.00 | False |
| R4 | 2 | R2 | -0.25 ≤ 3.13 ≤ 5.75 | True |
| R4 | 3 | R1 | 2.25 ≤ 7.10 ≤ 8.00 | True |
| R5 | 1 | R4 | -10.75 ≤ 18.70 ≤ -3.75 | False |

Check 2: Compare all successive pairs of records surrounding and outwards from the current record (*i*)

$$\left(E_{i+y}^{min} - E_{i-y}^{max}\right) - 2 \le \left(A_{i+y} - A_{i-y}\right)/30 \le \left(E_{i+y}^{max} - E_{i-y}^{min}\right) + 2$$

| *i* | *y* | *i + y* | *i - y* | Evaluated Equation | Result |
|---|---|---|---|---|---|
| R2 | 1 | R3 | R1 | -4.75 ≤ 6.13 ≤ 1.00 | False |
| R3 | 1 | R4 | R2 | -0.25 ≤ 3.13 ≤ 5.75 | True |
| R3 | 2 | R5 | R1 | -6.50 ≤ 25.80 ≤ 2.25 | False |
| R4 | 1 | R5 | R3 | -3.75 ≤ 19.67 ≤ 3.25 | False |
| R4 | 2 | R6 | R2 | -6.25 ≤ 23.23 ≤ 1.50 | False |
| R5 | 1 | R6 | R4 | -8.00 ≤ 20.10 ≤ -2.25 | False |

Start of new episode (R5)

3 **Assign start of new episode if all comparisons from both checks are false**

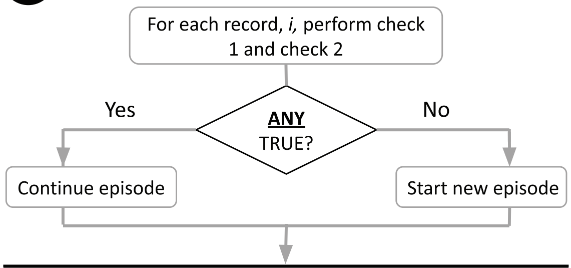


| Episode Number | Start Date | End Date |
|---|---|---|
| 1 | 2019-11-01 | 2020-07-01 |
| 2 | 2022-01-13 | 2022-02-24 |

**Figure S1. Detailed steps of Pregnancy Progression Signature (PPS) Algorithm using a fictional example patient.** 1) The list of 74 concepts (Table S3) was used to search for signatures of progressing pregnancy concepts across each patient record using an adaptation of longest increasing consecutive subsequence analysis. Outlined in red is a record that is discordant with the other records. 2) Time intervals across these concepts in the patient data were compared to their expected intervals in terms of gestational month assigned by clinicians (Table S3) in order to build plausible pregnancy episodes, where i is the current gestational timing record in the patient data, A is the actual (observed) patient record date, E is the expected concept gestational timing in months, x is the iteration across records for check 1 and y is the iteration across records for check 2. Two additional months were added to the expected ranges in order to allow for a margin of error in when the concepts were recorded in the data. Outlined in purple is the record that indicates the start of a new episode. 3) Upon checking each record, i, if any records comparison from check 1 or check 2 evaluate to a true result, the episode is continued and it is assumed the patient still is progressing with the same pregnancy, i.e. the actual difference in dates matches the expected difference in dates based on the concepts gestational time ranges. If none of the checks evaluate to true and there are > 2 months (minimum retry period for any pregnancy outcome) between consecutive record dates, a new episode begins. The large number of comparisons performed per date in the patient records was due to historical records in the patient data intervening with the true progressing sequence of pregnancy gestational timing concepts, meaning that concept comparisons often had to be skipped and instead surrounding concepts were assessed to determine whether to continue an episode or start a new one.

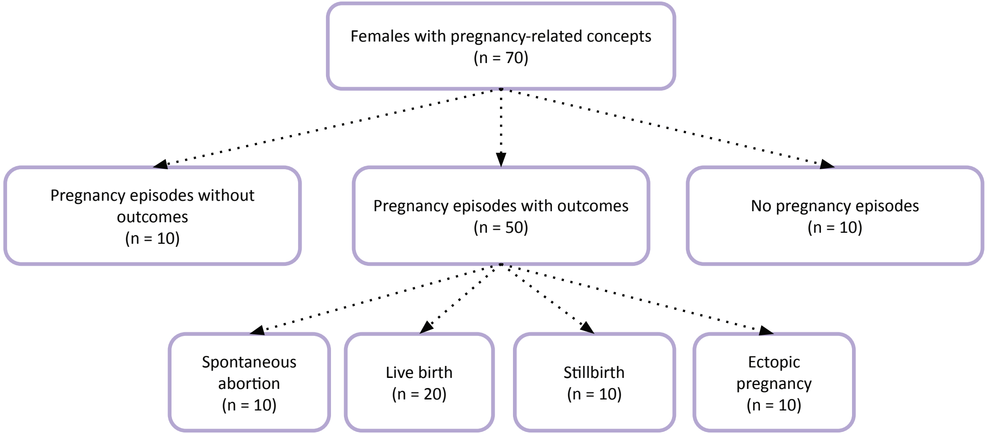


**Figure S2. Selection of patients used for independent clinician validation.**
* As these cases were randomly selected for clinician validation they do not reveal information about the subpopulation and are not subject to the same policy regarding obfuscating small numbers of patients.

| Concept category | Count and proportion of sites (out of 72) with at least one person matching the concept | | Count and proportion of persons (out of 633,914 across all sites) matching the concept | | Count and proportion of unique persons who have the concept recorded but who are missing corresponding values | |
|---|---|---|---|---|---|---|
| Gestation period, X weeks | 70 | 97.2% | 499,655 | 78.8% | 0 | 0.0% |
| Gestational age | 22 | 30.6% | 20,184 | 3.2% | 11,189 | 55.4% |
| Last menstrual period start date | 21 | 29.2% | 22,158 | 3.5% | 16,532 | 74.6% |
| Delivery date Estimated | 19 | 26.4% | 17,280 | 2.7% | 10,217 | 59.1% |
| Gestational age in weeks | 19 | 26.4% | 10,761 | 1.7% | 2,818 | 26.2% |
| Gestational age in days | 19 | 26.4% | 6,014 | 0.9% | 495 | 8.2% |
| Gestational age Estimated | 18 | 25.0% | 10,352 | 1.6% | 381 | 3.7% |
| Gestational age Estimated from conception date | 15 | 20.8% | 4,980 | 0.8% | 4,980 | 100.0% |
| Gestational age US composite estimate | 10 | 13.9% | 5,839 | 0.9% | 5,687 | 97.4% |
| Delivery date Estimated from last menstrual period | 9 | 12.5% | 3,790 | 0.6% | 3,229 | 85.2% |
| Delivery date US composite estimate | 9 | 12.5% | 4,010 | 0.6% | 3,108 | 77.5% |
| Gestational age Estimated from last menstrual period | 8 | 11.1% | 4,187 | 0.7% | 3,775 | 90.2% |
| Estimated date of delivery | 5 | 6.9% | 8,776 | 1.4% | 8,776 | 100.0% |
| Date of gestational age estimate | 2 | 2.8% | 3,022 | 0.5% | 3,021 | 100.0% |
| Gestational age Estimated from physical exam | 1 | 1.4% | 3,050 | 0.5% | 3,050 | 100.0% |
| Date of last menstrual period | 1 | 1.4% | 365 | 0.1% | 363 | 99.5% |
| Length of gestation at birth | 1 | 1.4% | 11,317 | 1.8% | 34 | 0.3% |

**Figure S3. Count and proportion of concepts by persons and sites.**

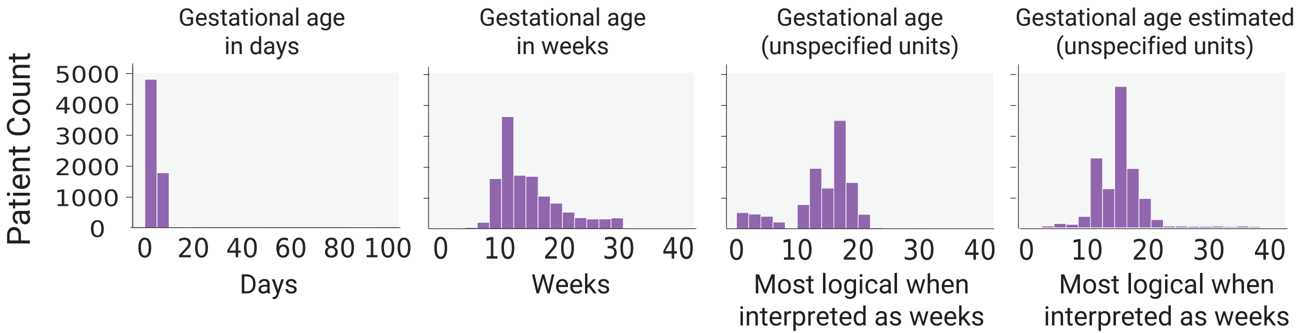


**Figure S4. Distribution of values of gestational aging concepts in the initial reference cohort.**

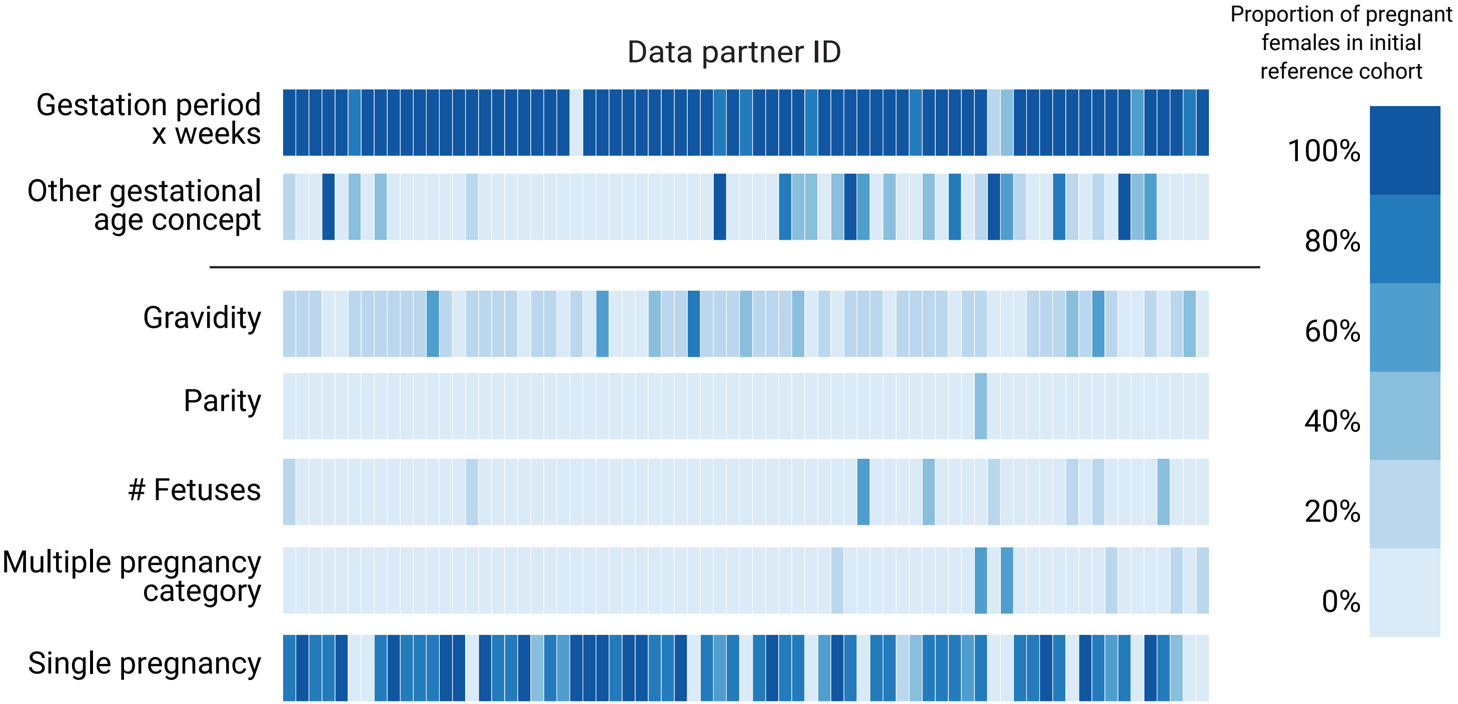


**Figure S5. Heatmap of proportion of pregnant persons in initial reference cohort by site with concept for Gravidity, Parity, Single Pregnancy, Multiple Pregnancy, Gestation Period, X weeks, Other Gestational, and Number of Fetuses**

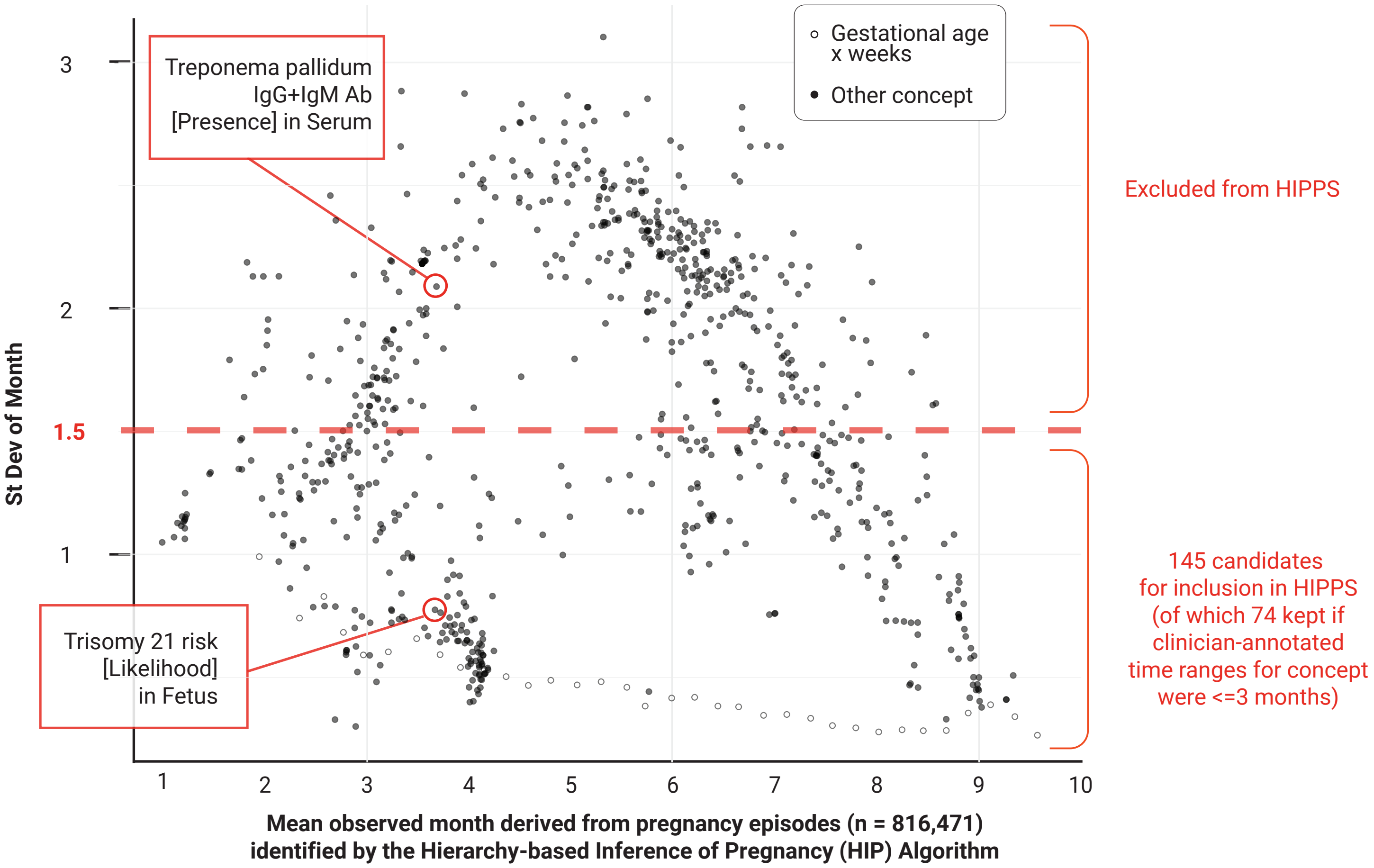


**Figure S6. Empirical estimation of threshold for gestational timing specificity for concepts,** with standard deviation of mean month in gestational timing vs average month the concept occurs during pregnancy (for any outcome category) as an indicator of specificity. Two concepts are called out as examples that were excluded and included in the HIPPS approach.

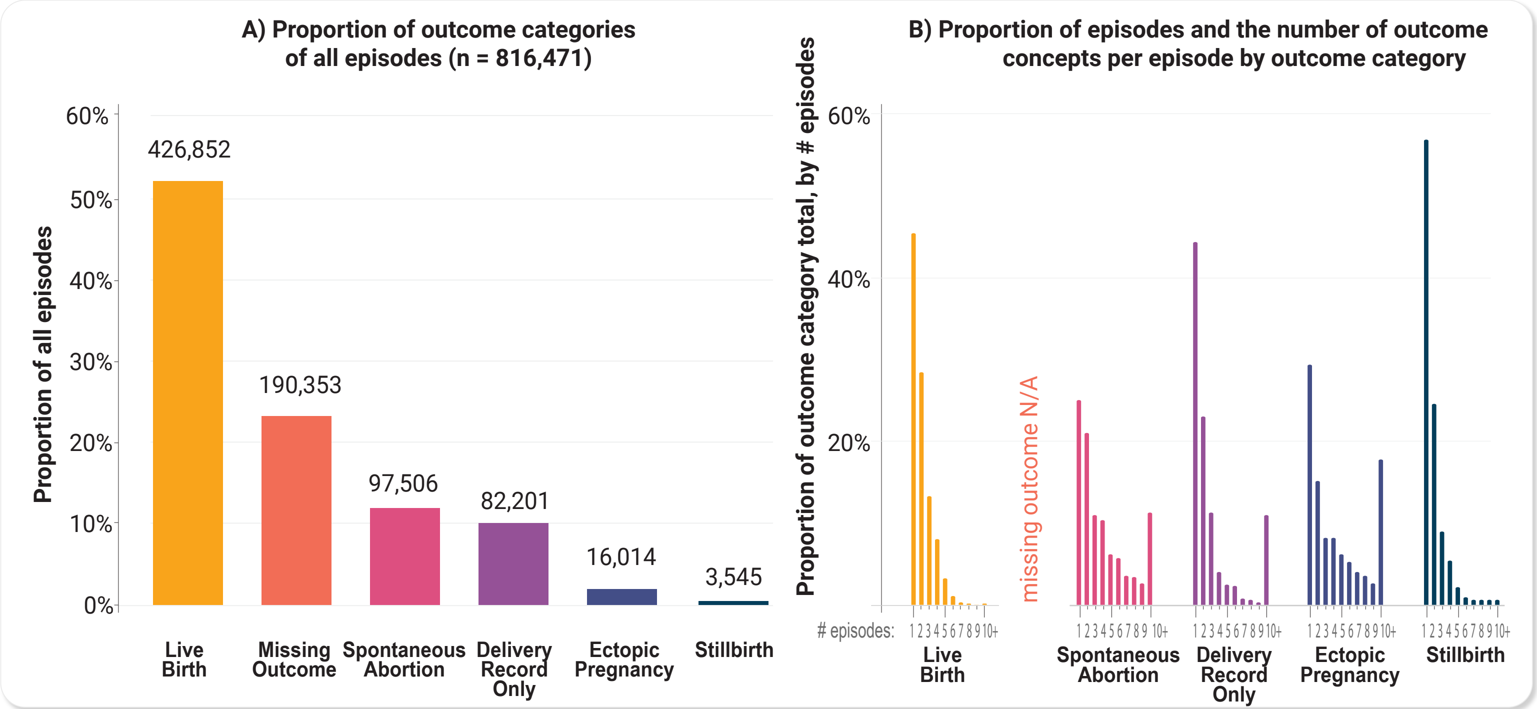


**Figure S7. A) Histogram of the number of outcome concepts per episode by outcome category. B) Histogram of episodes with week-level resolution only (N=564,762) by outcome category.** Number of outcome concepts were determined from the outcome date to 28 days after.

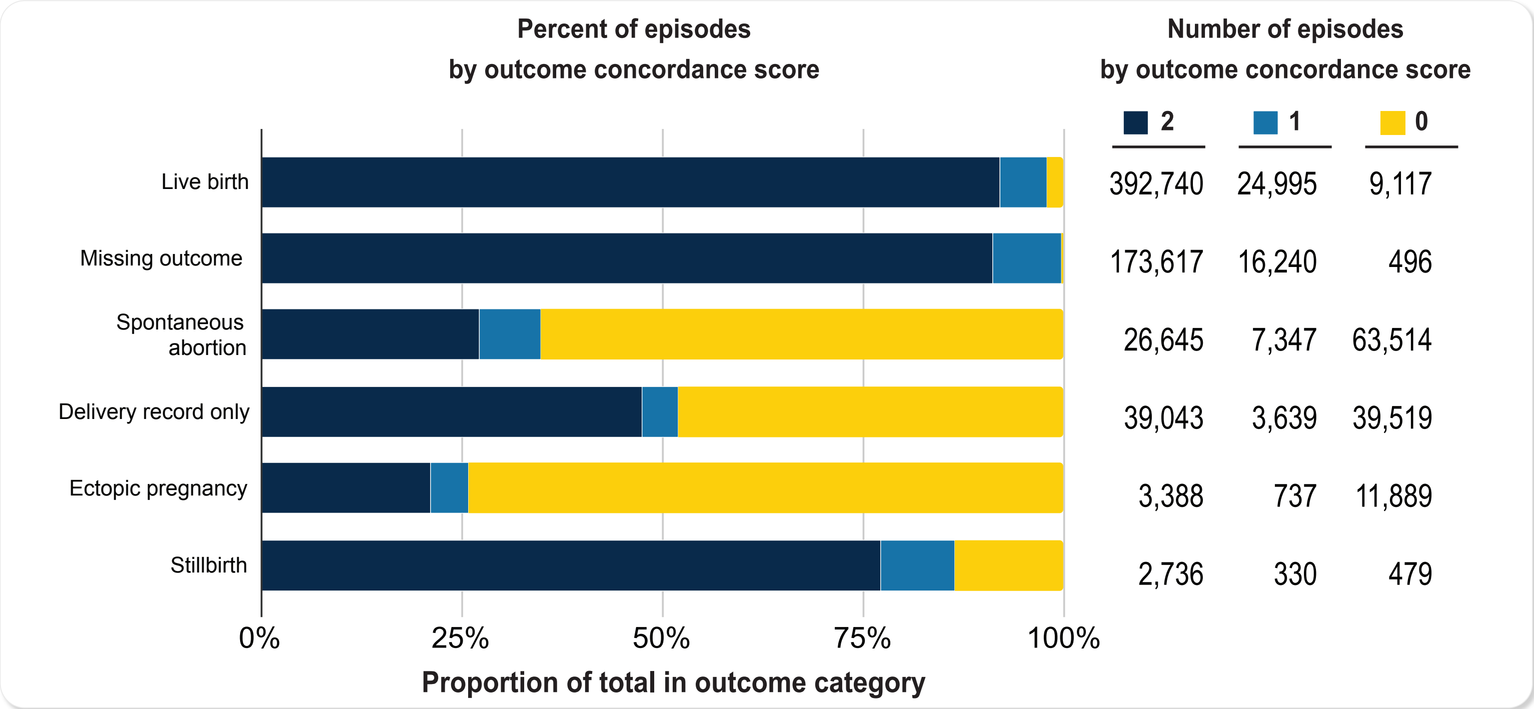


**Figure S8. Outcome concordance scores by outcome category.** An outcome concordance score of 2 has an outcome within the expected term duration and is supported by both HIP and PPS. An outcome concordance score of 1 has an outcome within the expected term duration. An outcome concordance score of 0 does not have an outcome within the expected term duration.
